# Public Involvement in Complex Theorising – A co-produced logic model of the role of context in shaping child health

**DOI:** 10.1101/2025.03.10.25323571

**Authors:** Dylan Kneale, Alison O’Mara-Eves, Bridget Candy, Lizzie Cain, Jessica Catchpole, Angela Chesworth, Sandy Oliver, Katy Sutcliffe, Niccola Hutchinson Pascal, James Thomas

## Abstract

**Introduction:** There is increasing work towards drawing on theory, implementing co-production, and accounting for complexity within the production of systematic reviews for public health. In this paper, we report on the process of co-producing a theory, in this case a graphical articulation of theory in the form of a logic model, which describes how contextual factors influence children’s health.

**Methods:** We undertook a series of three online co-production workshops involving 18-20 participants in each, and worked with an advisory group of experts with professional and lived expertise. An online virtual whiteboard was used to support the identification of factors that contributed to poorer childhood health, explanations for these factors, and connections between different factors.

**Results:** We initially focussed our work on childhood obesity. However, co-production was transformational in switching the focus of the logic model away from a narrow focus on Body Mass Index as a measure of obesity, to a more holistic theory of factors that shape children’s health, recognised as the intersection between healthy eating, physical activity and mental wellbeing. Theorising with a diverse range of co-producers helped us to recognise the stigmatising impacts that an exclusive focus on clinical measures of children’s health can have, and the way that a narrow clinical focus inhibits theorising the complexity and drivers of poorer health.

**Conclusion:** Co-production led to a switch in theorising away from narratives of children’s health that focus closely on personal responsibility, towards narratives that explore structural and contextual drivers of health.

**Patient or Public Contribution:** The logic model was entirely driven by the contributions of researchers, those with lived experience (e.g. as parents and/or who have experienced poor health), and those with professional experience (e.g. as teachers) who worked together to co-produce the model. An advisory group composed of people with a similar range of expertise helped to shape the conduct of co-production and dissemination (including in the preparation of this manuscript).

## 1.1 Introduction

Theory should underpin all forms of research, and the role of theory is becoming increasingly recognised within evidence synthesis ^1,2^. Common across theories – be they implicit or explicit ^2^, grand or low-level ^1^ – is that a theory is itself a product of synthesising knowledge, that involves articulating different concepts and how they relate to each other, which can then be used to generalise to different scenarios beyond the immediate ^2^. However, different forms of knowledge are valued unequally, and historically knowledge gained through lived experience of a health condition or service has been overlooked as a knowledge source in the development of theory ^3^. In this research, we report on the process and results of research where a theory around the influencers on children’s health was co-produced.

Logic models are a graphical approach to visualising and/or creating theories around how an intervention activates a series of processes that are thought to create change ^4^. A broad distinction can be made between process-based logic models, which support theorising of the granular steps taking place within an intervention or service, and systems-based logic models. The latter involve greater theorising of the relationship between an intervention and its context than the former ^5,6^.

Alongside a trend towards clearer articulation of theory (and particularly logic models ^7^), two other trends are directing evidence synthesis methodology. The first of these is a recognition of the value of public and patient involvement (PPI) in the creation of evidence syntheses ^8^. In particular, co-production is recognised as a way of transforming the relationship and power differentials between researchers and non-researchers to enable more equitable decision-making ^3^. Co-production can be hugely beneficial to the quality of research, although these benefits are challenging to quantify ^3^. The second trend is a movement towards recognising and unpacking the role of complexity within evidence syntheses, and particularly around the wider contexts within which interventions are implemented ^9^.

However, understanding which specific features of context may influence health outcomes is often not fully understood and is under-theorised ^10^.

In this research, we integrated these three elements (logic models, PPI, and unpacking complexity) within a project that sought to co-develop a theory (a systems-based logic model) around children’s health (and initially, specifically around obesity) that was focussed on identifying which contextual features harmed or promoted children’s health.

### 1.1 Childhood obesity and theorising factors that drive childhood obesity

Childhood obesity has multiple negative physical and psychological health implications ^11^. The global prevalence of obesity has increased over the last decades and is set to continue ^12^. Theoretic frameworks to understand the causal mechanisms and context of childhood obesity have historically focused on individual behaviour-change such as the Theory of Planned Behaviour ^13^. Alongside recognition of the broad context of influences on obesity, whole system approaches are gaining greater traction. Such approaches in the context of obesity are a way of considering how individuals, groups, services and organisations interconnect and influence each other ^13^. They allow consideration of both immediate and more distal contextual factors including for example industry, and government policy. They conceptualise the drivers of obesity as a system with a collection of interdependent parts, highlighting how influences from one part of the system can affect another. Within this conceptualisation, obesity is not necessarily the ‘outcome’ of a chain of potentially modifiable factors, but a single component of a large dynamic system; and it is the interplay of a complex web of interacting relationships that gives rise to what is observed as obesity. This conceptualisation then highlights the = complexity of what needs to be done to reverse trends. One such whole system approach is the UK government’s Foresight Obesity report ^14^, a key output of which was a map of drivers theorised to influence (adult) obesity, which is seen as a turning point where obesity came to be understood in terms of whole systems complexity ^15^.

The Foresight obesity systems map visually sets out both the complexity of the drivers of obesity which fell within the responsibility of multiple agents (e.g. individual, commercial, voluntary sectors and government) ^16,17^. There are 108 drivers of obesity included in the Foresight Systems map, that are divided into seven interlinking areas of influence on a person’s energy intake: individual biology; individual activity; environmental activity; individual psychology; societal influences; food consumption; and food production. All these link to individual’s energy balance placed in the centre of the system. What visually dominates the systems map is the over 300 lines that connect the factors, alongside feedback causal loops of different sizes dependent on likely strength of influence.

The Foresight model is of interest here as it takes a systems perspective and provides a theory of influencers on obesity. However, it was based on the input of experts, who were asked what they believe to be important and who collectively published thirty-four reviews and opinions pieces that contributed to the development of the model ^18^. ‘Non-scientific’ input included representatives from government departments, as well as the retailer Tesco ^15^.

Crucially, the development of the model did not involve people with lived experience. Moreover, it was not specific to the focus of this work - examining childhood obesity, which plausibly has very different factors and pathways to a general or adult-focused model.

## 2 Methods

### 2.1 Study Aims

The aims of this study were to:

i. develop a systems-based logic model of the factors that influence children’s capacity to maintain a healthy weight;
ii. to understand what happens when a logic model is co-produced through the input of a range of experts – those with lived expertise (as parents, children or carers), those with professional expertise (as teachers or health professionals) and those with academic expertise (as researchers in obesity or childhood).

The logic model was to focus on the context surrounding interventions that are implemented in schools. The logic model was to be used to inform a larger project on evidence synthesis methods (not reported here) that involved the development of novel meta-analysis methods. These new meta-analytic methods were intended to support the exploration of context within systematic review evidence, but were not co-produced. However, the selection of which contextual factors to explore in the meta-analytic methods was intended to be grounded in theory (i.e., derived from the co-produced systems-based logic model).

### 2.2 Setting and Context

During the midst of the pandemic in 2020, and with evidence showing that obesity was linked with a worse COVID-19 prognosis ^19^, the government launched a new strategy for child and adult obesity ^20^. This was the fourteenth obesity strategy launched in the past three decades ^21^. While the strategy was a marked departure from several previous strategies in containing a greater number of restrictive policies that sought to change the availability of unhealthy foods (e.g. banning price promotions around unhealthy products) ^21^, not all of these policies were actually implemented ^22^. Notably, the strategy did not include policies targeted solely at physical activity ^21^, but did contain policies targeting diet alone, or diet in conjunction with physical activity.

The project took place between 2021-2022, at a time when COVID-19 pandemic restrictions were gradually being lifted in the UK and the COVID-19 vaccine programme was being implemented. Restrictions on social gatherings were still in place for part of this work, and all of the work took place remotely, with the academic research team neither meeting with one another nor with new co-producing colleagues. The whole project – co-production of a logic model and development of meta-analytic methods – was conducted within a rapid timeframe of ten months.

### 2.3 The academic research team and advisory group

Within the [ANONYMISED]-based research team we brought a range of experiences including around evidence synthesis, public health, children’s participation and rights, weight management research and experiences, and around participatory methods. However, to support the co-production of the model, we also worked in partnership with members of the Co-Production Collective, who were instrumental in training us on the values, methods, and processes of co-production (see ^3^).

To support the research, we assembled an Advisory Group (AG) of experts with lived and professional experience. We approached membership and management of this Advisory Group through the principles of co-production, although the AG was not directly involved in co-producing the logic model.

In total, nine people agreed to join the advisory group including academics (3), public health practitioners (2), teachers (1) and people with lived experience (2). The advisory group was given email updates and opportunities to comment throughout the project, and met (virtually) three times. All meetings were hosted on Zoom. The first advisory group session involved informing the group of the purpose of the study, providing an opportunity for members to get to know each other, clarifying expectations for the research, and determining how to organise the co-production of the logic model. The second advisory group meeting occurred after the stages of co-production to discuss the draft logic model and its implications for future work. The third advisory group meeting occurred after a working draft of the model had been developed, and when we had progressed to using the model to inform the development of meta-analytic methods. A fourth meeting had been planned, but in the end, it was deemed that final inputs would be best gained through email contact. This was in part due to the timeframe of the work, which was already tight but further compounded with the emergence of a new coronavirus variant placing strain on people’s time.

One crucial way in which the AG strengthened the research, and our approach to the research, was in advising the team on how to have conversations about sensitive topics like obesity when co-producing the logic model. While the AG members were intended to occupy a role that was exterior to the co-production team, we came to regard them, and particularly both AG members with lived experience, as integral to the project and as co-producers and co-authors of this article.

### 2.4 Forming a team for co-producing a systems-based logic model

To co-produce a systems-based logic model, we planned a series of online workshops. The co-production workshops were designed to gain insights of local factors relating to child health. There were two initial workshops to develop the model, then a third workshop with a broader audience to check and challenge the model. The workshops were conducted virtually and hosted on Zoom and were scheduled to last two hours.

#### 2.4.1 Co-production Workshops 1 and 2

##### Focus

The first workshop was intended to identify important concepts, and the second workshop was intended to identify the relationships between these concepts.

##### Recruitment

Recruitment of co-producers took place informally through a mixture of targeted and open recruitment. Targeted recruitment drew on the personal and professional links of the academic research team and the Co-production Collective. Open recruitment was through mailouts, blogs hosted on the Co-production Collective website, and a website set up for the project. Remuneration for people’s time was offered based on established guidance which reflected attendance at the workshops plus any preparation or follow-up work ^23^.

In Workshop 1, in addition to the project team of academic researchers and two members of the Co-Production Collective (that is, the [ANONYMISED]-based co-producers), there were 11 co-producers, who described their relevant expertise primarily as: GP and advocacy (n = 1); teaching (n = 4); lived experience (n = 4); and research (n = 2).

In Workshop 2, we invited the same people who were invited to Workshop 1. In addition to the [ANONYMISED]-based co-producers, there were 13 attendees, who described their relevant expertise primarily as: GP and advocacy (n = 1); teaching (n = 3); lived experience (n = 5); research (n = 2); occupational therapy (n = 1); and nutrition and advocacy (n = 1). A member of Co-Production Collective facilitated both workshops and liaised with co-producers before and after the events. All co-producers were aged 18+.

##### Process

The first two workshops followed a similar pattern. Colleagues from the Co-production Collective who were experienced in facilitating dialogue between diverse groups of people laid out the expectations for how we would work together, including reviewing the principles of co-production (see ^3^). Workshops 1 and 2 then started with a brief introduction to the project and its goals, followed by discussions in three breakout groups with a facilitator from the project team in each of the breakout groups. The discussions were guided by starter questions (see table 1, below). A second project team member posted notes of the points raised on a live, virtual whiteboard).

**Table 1:**
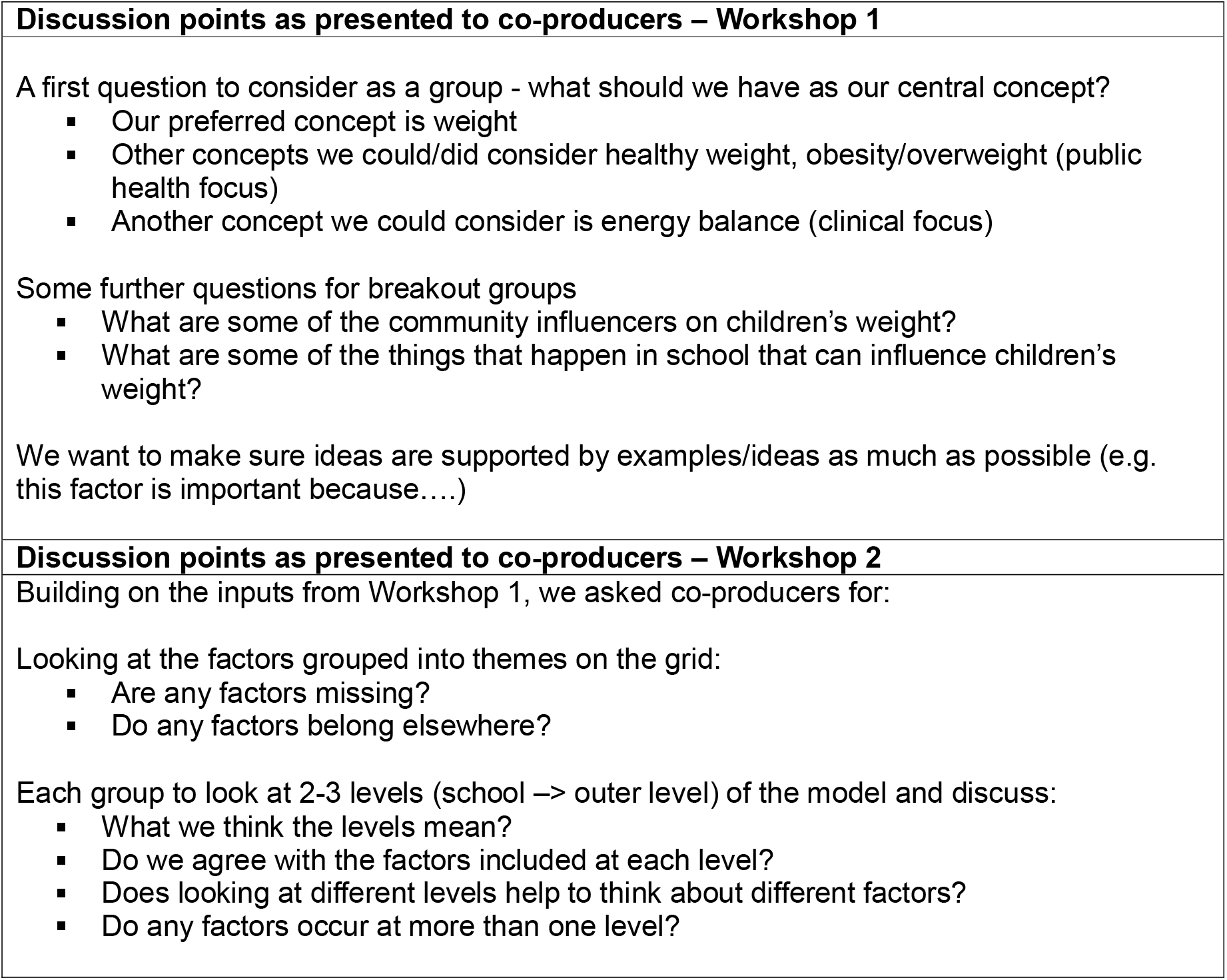
Discussion points presented to co-producers across co-production workshops 1 and 2.

To facilitate the discussion in workshop 1, we drew on the ‘domains’ identified in the Obesity Foresight map as a starting point, and expanded on these, and asked people to consider factors relating to the following domains: (i) food; (ii) biological factors (explained as things to do with how the body works including genetics and metabolism); (iii) social factors (which could be personal, community and cultural factors); (iv) activity and behavioural factors; (v) developmental factors (things to do with age transitions); (vi) economic factors (things to do with spending and earning money); (vii) infrastructure and environmental factors (things around us in our daily lives such as transport and buildings); (ix) psychological factors; (x) media. We also added (xi) medical factors (things to do with illness and injuries including the side effects of medications); (xii) school factors (what happens in school); and (xii) space for co-producers to identify ‘other’ factors that did not fit into the other categories.

One element that the [ANONYMISED]-based co-producers attempted to convey was that much of what might be represented within the model might not be captured within studies or were not easy to measure in practice, as people’s experiences are often more complex than are represented in the available data. However, the [ANONYMISED]-based co-producers also emphasised that this did not belie or detract from the importance of identifying these factors. This was part of efforts to be clear about the expectations of how the model was going to be used initially within the project.

The workshops were spaced three weeks apart to keep the momentum and to try to ensure attendance at both. Between the first and second workshop, two of the [ANONYMISED]-based co-producers (DK and AOME) attempted to group the virtual post-it notes that had been recorded and consolidate any duplicates from the break-out groups. In grouping the notes, it was also identified that the factors represented on the post-its also fell within different ‘ecological’ levels of influence. We used an adapted social-ecological model ^3^ as the basis for organising the post-it notes subsequently. The post-its were grouped across the following levels: (i) individual; (ii) household, family, and friends; (iii) school; (iv) neighbourhood (place-based); (v) cultural community (incl. social media); (vi) economic systems; (vii) socio-political, infrastructure, national policy, media; (viii) cross-cutting factors (temporal and historical trends).

The arrangement of the different factors identified in Workshop 1, the configuration of the outcome, and the arrangement of factors across the different levels of influence became the main sources of discussion in Workshop 2.

#### 2.4.1 Co-production Workshop 3

##### Focus

Once the themes/concepts were organised, we needed a visualisation method that: preserved the levels and concepts; could represent concepts ‘hierarchically’ (from more to less detailed); could represent some ‘logical’ relationships; and could preserve the original sentiments for reference. We continued to use Miro for this visual representation. The aim of workshop 3 was to check and challenge the logic model, as well as introduce new methodological developments.

##### Recruitment

We held a third workshop in which we invited the original participants back, plus a new group of policymakers and practitioners. Workshop 3 had 12 attendees: 7 ‘returners’ and 5 new attendees recruited through our networks.

##### Process

Participants were provided with a link to the interactive model and a short instructional video prior to the workshop. During the 2-hour workshop, we presented the project progress and then used breakout groups to seek feedback from the co-production team and the wider set of participants.

None of the co-production workshops were recorded to enable co-producers to speak more freely about their experiences and to voice their opinions.

### 2.5 Study data

The results reported here are based on:

1. An analysis of the features of the co-produced logic model
2. Summaries of informal reflective discussions among [anonymised]-based co-producers that took place after co-production workshops and within advisory group meetings
3. Summaries of reflections submitted by co-producers based outside [ANONYMISED], who were were invited to submit their reflections to the Co-Production Collective. These were summarised by LC.

These helped us to identify the distinctive contribution that co-production made to the development of the logic model.

### 2.6 Ethics

This research followed the Economic and Social Research Council’s research ethics framework. Ethical approval was gained from the [anonymised] (REC 1498). In line with institutional policy at the time, informed consent was sought from co-producers based outside [ANONYMISED] for participation in the workshops, although we acknowledge that obtaining consent is not usually necessary in cases where co-producers are equal partners in the research.

## 3 Results

The aim of the study was to develop a systems-based logic model of the factors that influence children’s capacity to maintain a healthy weight. The following sections describe the product of this work and the extent to which this aim was met. For the most part, due to the fast-paced nature of the co-production workshops, it is difficult to disentangle the sources of the different contributions to the model. Below, however, we highlight key points in which the unique contributions of the collaborators from beyond the core research team were particularly evident in developing the logic model.

### 3.1 The switch in focus from childhood obesity to children’s health

Much of the discussion in the first workshop centred around the outcome that should be the central focus of the logic model. The [ANONYMISED]-based co-producers had used childhood obesity as a starting point, and based on previous experience of conducting reviews in this area had anticipated that measures of weight (and particularly Body Mass Index (BMI)) would predominate as the primary outcome in the intervention literature. Co-producers were unanimous in questioning the value of the focus on children’s BMI. Co-producers with lived experience of obesity or childhood obesity and parents voiced concerns that obesity was a hugely stigmatising condition for children, and that this stigma was as damaging to the transition to adulthood as the health implications of obesity—and likely much more so. A particular concern to one co-producer was the ‘malleability’ of BMI to the influence of public health interventions. Among health practitioners in the group, it was felt that standardised measures of weight such as BMI could be useful in decision-making, although had their limitations. For public health practitioners and researchers, BMI as a measure was recognised as a narrow construct and was not a useful reflection of the complexity of children’s health. Further, there was no support for energy balance as a primary outcome of the model, despite this being the central focus of the original Foresight Obesity model.

During the first co-production workshop, therefore, it was agreed that the outcome central to the logic model should reflect a broader conceptualisation of children’s health than BMI. Much of the discussion, reflecting the expertise in the room and the framework used, continued to focus on influences on children’s diet and physical activity and factors influencing childhood mental health. Between the first and second workshop, the [ANONYMISED]-based co-producers aimed to seek clarification around the parameters of the broader outcome and proposed in the second workshop that children’s health should be operationalised in the model as the intersection between healthy eating, physical activity, and mental health (see Figure 1). While diet and physical activity are often co-occurring as the foci of multicomponent weight management interventions ^24^; mental health is often treated only as a secondary outcome or neglected altogether. In addition, healthy eating and physical activity (HEPA) were already recognised as co-occurring in some strategies; the addition of mental wellbeing (creating the acronym HEPAM) was viewed as reflecting the concerns of all co-producers that issues around stigma and weight were underappreciated within debate within this space.

**Figure 1:**
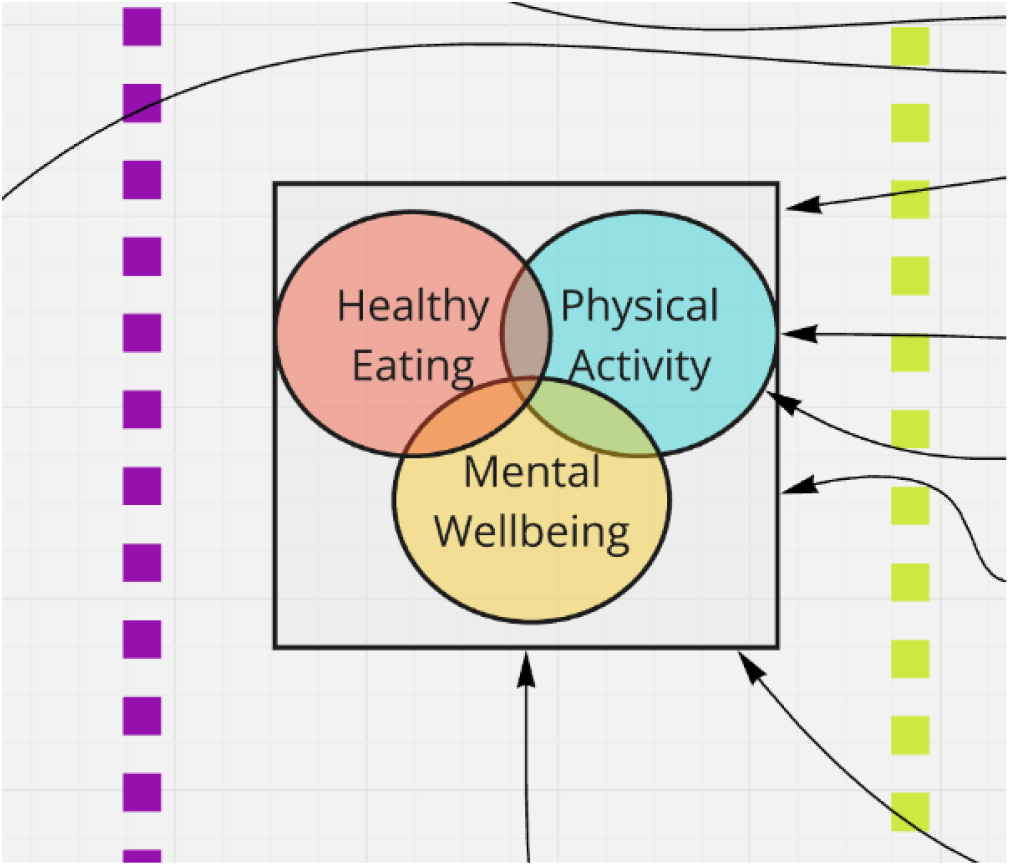
Section of logic model (see explanation in text)

The focus on HEPAM – where good child health is determined through the intersection of healthy eating, physical activity and mental wellbeing - was viewed by almost all co-producers as a satisfactory way of attempting to capture the complexity of the discussions within the first workshop. They were viewed as joint outcomes that school-based interventions could theoretically address. However, for a very small number of co-producers, the switch in outcome from initial proposals to examine BMI to HEPAM was not transformational enough in addressing concerns around the damaging impacts of labelling children as obese, or likewise, as ‘unhealthy’.

### 3.2 An overview of a systems-based logic model of children’s health

The central outcome of the logic model is a broad conceptualisation of children’s health that reflects an intersection between healthy eating, [partaking in] physical activity, and mental well-being. The model is organised according to two central principles – that influencers on children’s health can be grouped into broad domains (i.e., different types of factors) and that influencers on children’s health occur at different socioecological levels; these are described below. Due to its complexity, the model is not reproduced here, although the link to the model is available <u>here</u>, and a link to a video to explain how the model can be used is available <u>here</u>.

Figure 2 below shows part of the model, which focusses on cultural community factors and elements around food as a domain. Here we can see the main factor (food culture) and a number of sub-factors relating to food culture. Each circle with the letter ‘D’ indicates that further text providing an explanation is available; in the example below, the social side of food and the reality that going to food outlets is a social/cheap activity is used to support ‘food as a social activity’. The model here recognises that school-based interventions to improve children’s health can be implemented in contexts where eating out is a social activity of significance.

**Figure 2:**
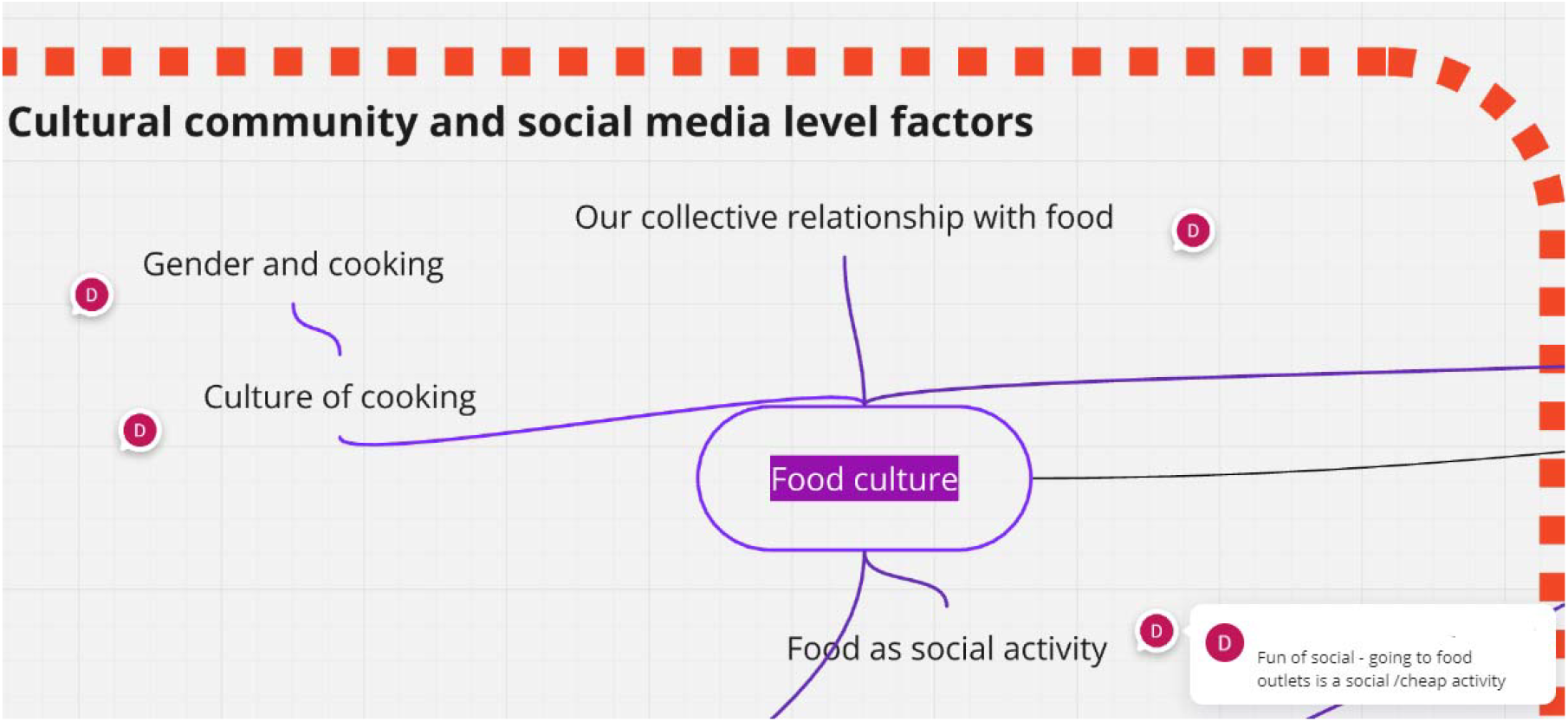
Section of logic model focusing on food culture themes (see explanation in text)

To aid the development of the theory, factors influencing children’s health were organised according to broad domains (see methods). These domains are colour coded, and more granular models that show both the factors (core concepts) and subfactors (granular details) belonging to a domain were also created as separate models (see table 2 for an overview). For example, ‘how children spend their time’ is a main factor in the ‘activity’ domain, but several subfactors of ‘how children spend their time’ (i.e., more granular descriptions) are also represented in the model, for example ‘screen time’ or children’s ‘routines’ or children’s ‘play’. These subfactors are supported by a description that provides an approximation of why the co-production team determined the factor an important influencer on children’s health in the context of school-based interventions. In the case of ‘play’ for example, the emphasis was on children’s ability to play freely and not only through organised sports.

**Table 2:** A description of the contents of the different domains within the logic model (see text for further information)

| Domain name | Example strand | Number of factors | Number of sub-factors | Number of levels in which the domain is represented |
| --- | --- | --- | --- | --- |
| **Psychological** | Children's agency <SF> Household dynamics -> Children's psychology -> Motivation for Healthy Behaviours -> HEPAM | 14 | 11 | 5 |
| **Infrastructure and Environment** | Access to leisure resources -> Children's environment -> Capability to be healthy -> How children spend their time -> HEPAM | 4 | 3 | 3 |
| **Media** | Social media -> Motivation for healthy behaviours -> HEPAM | 3 | 2 | 1 |
| **Biological and medical** | Interaction between genes and environment <SF><br>Genetics -> HEPAM | 3 | 1 | 1 |
| **Activity** | Screen time <SF>How children spend their time -> HEPAM | 8 | 6 | 6 |
| **Schools** | Policies around how children spend their time -><br>How children spend their time within school -> HEPAM | 6 | 6 | 2 |
| **Developmental** | Puberty -> HEPAM | 2 | 1 | 1 |
| **Economic** | Globalisation <SF> Macroeconomic policy and values -> Poverty -> HEPAM | 9 | 4 | 3 |
| **Food** | Household attitudes to food <SF> Child attitudes to food -> [Capacity to make] Informed choices -> What we eat and drink -> HEPAM | 15 | 24 | 7 |
| **Other** | Evidence ecosystem (how evidence is used) | 7 | 15 | 3 |
|  | **Total** | 71 | 73 | 8 (including cross-cutting trends) |
| **Key** -> = Influences; <SF> = Sub-factor/feature of |  |  |  |  |

These factors can also occur at different points in children’s lives. For example, ‘how children spend their time’ is a factor that is influenced by, and occurs within, schools, neighbourhoods, culture, economic systems, households and families, and by factors occurring at the individual-level. A number of the levels and factors are clearly beyond the purview of any individual policy, service, or intervention to improve children’s health (for example, a school-based intervention to improve children’s health could not influence broad culture). They may, however, be important factors to consider in interpreting evidence, and particularly in interpreting results across different settings.

### 3.3 Personal choice vs Upstream Determinants

A noteworthy feature of the model is its focus on upstream determinants. Co-producers questioned the extent to which individual factors reflected genuine ‘choices’ when individuals were making those choices in the shadow of large companies and inadequate policy responses. For example, co-producers described how the “food industry working actively against the driver - promoting idea of individual choice /blame” and questioned “morals in food industry”. In this respect, the model reflects the ‘commercial determinants of health’ that others have also cited as disproportionately responsible for poor public health outcomes ^24^. Such commercial determinants may be particularly important to consider in the case of children, who may have less control over their environments.

The model also includes a concern around the commercial determinants of health, and the role that online media plays in shaping children’s health. In fact, within the media domain, there was little mention of factors relating to broadcast media (TV or online streaming) and much more concern around social media and social networking as determinants of HEPAM. As may be expected in modern times, the model includes no mention of print media.

In line with discussions around the broad conceptualisation of children’s health, there was concern that there had been disproportionate focus in society and healthcare policy on individual and parental responsibility. A factor directly reflecting this around ‘the onus on individual not societal responsibility’ was included. Table 3 also shows that under a third of factors and subfactors named were placed in the individual or household, family and peer networks levels (29% (43/146)) with a far greater number placed in levels reflecting sociopolitical, economic and cultural and community levels (42% (61/146)). Co-producers also critiqued the language of policy around obesity, which routinely describes children being in the midst of a “war” or “crisis” or “epidemic”.

**Table 3:**
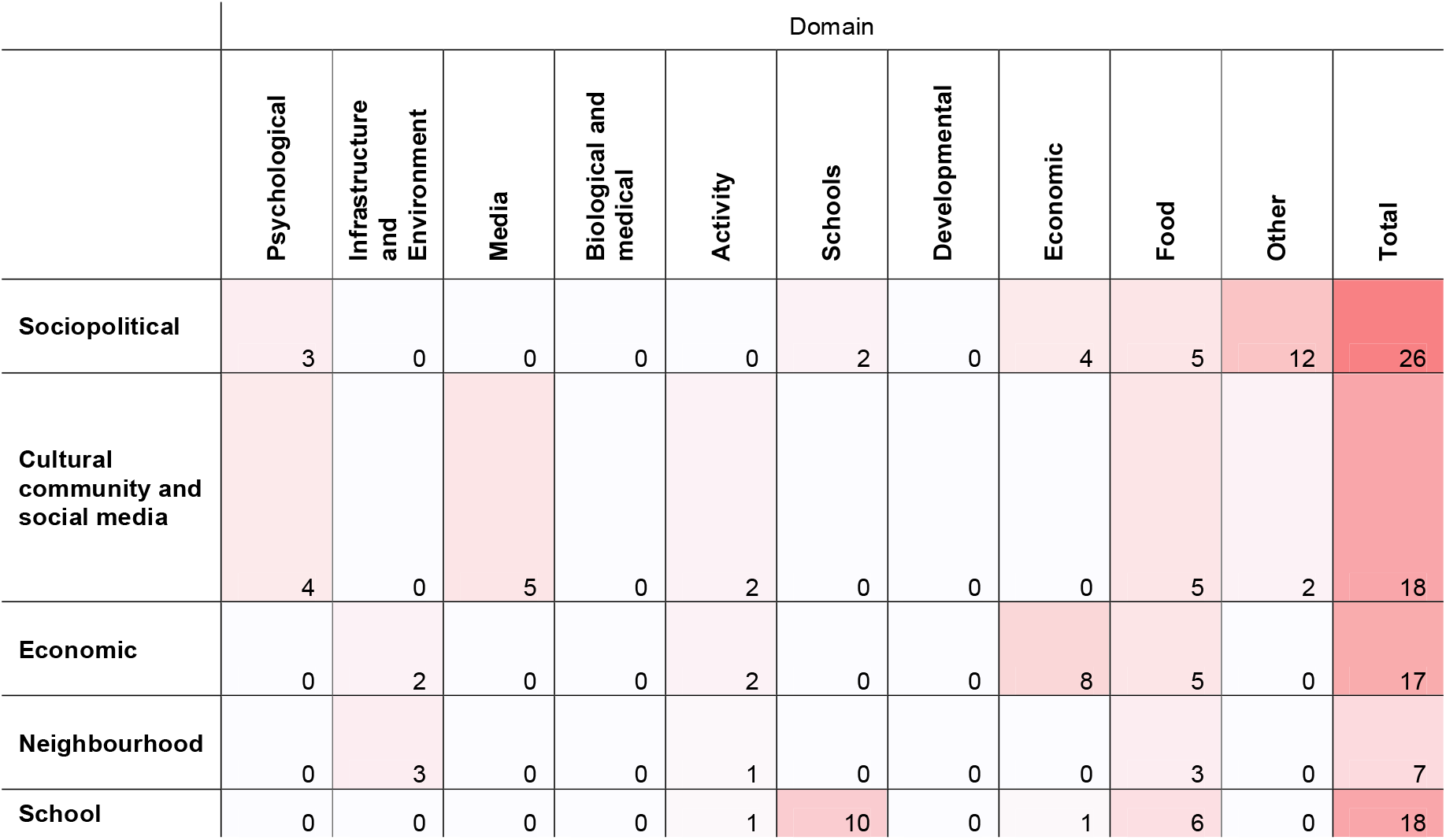
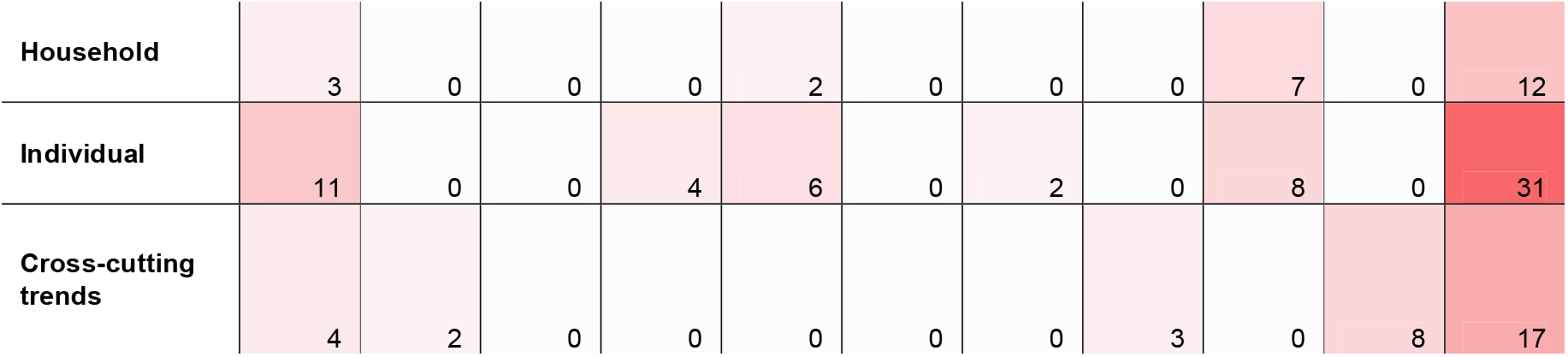
Number of factors related to each domain (psychological, media, etc.) represented in the different levels (individual, sociopolitical etc.) within the logic model (see text for further information)

|  | Domain |  |  |  |  |  |  |  |  |  |  |
| --- | --- | --- | --- | --- | --- | --- | --- | --- | --- | --- | --- |
|  | Psychological | Infrastructure and Environment | Media | Biological and medical | Activity | Schools | Developmental | Economic | Food | Other | Total |
| **Sociopolitical** | 3 | 0 | 0 | 0 | 0 | 2 | 0 | 4 | 5 | 12 | 26 |
| **Cultural community and social media** | 4 | 0 | 5 | 0 | 2 | 0 | 0 | 0 | 5 | 2 | 18 |
| **Economic** | 0 | 2 | 0 | 0 | 2 | 0 | 0 | 8 | 5 | 0 | 17 |
| **Neighbourhood** | 0 | 3 | 0 | 0 | 1 | 0 | 0 | 0 | 3 | 0 | 7 |
| **School** | 0 | 0 | 0 | 0 | 1 | 10 | 0 | 1 | 6 | 0 | 18 |
| **Household** | 3 | 0 | 0 | 0 | 2 | 0 | 0 | 0 | 7 | 0 | 12 |
| **Individual** | 11 | 0 | 0 | 4 | 6 | 0 | 2 | 0 | 8 | 0 | 31 |
| **Cross-cutting trends** | 4 | 2 | 0 | 0 | 0 | 0 | 0 | 3 | 0 | 8 | 17 |

However, despite being more orientated towards acknowledging commercial, social and political determinants of children’s health, some participants at the end of the third workshop indicated that environmental factors had still been downplayed, that mental health did not occupy a sufficiently prominent position, and that government choices such as austerity were not adequately represented.

### 3.4 What the model does not cover

As noted in the Methods, the first workshop was intended to identify different concepts and the second workshop was intended to identify the relationships between different concepts, but the second intention was not fully addressed. We simply ran out of time in the first workshop, in part reflective of budgetary and time constraints, and collectively agreed that it would be more fruitful to ensure that we had good coverage of key factors rather than moving onto look at relationships between an incomplete set of factors.

As a result, we have very few posited pathways between factors. Those that are included were those that were explicitly discussed in the meetings; there are likely to be others. In addition, the model does not tell us about the strength (magnitude) or any effects; the direction of any effects; any mediating or moderating factors; or whether any factors are supported by empirical evidence.

Finally, the model does not tell us about the relative importance of the different factors. As a part of a system, it is acknowledged that changes to any one factor in the system may lead to changes or adjustments in other parts of the system; the model does not make predictions about how any reactions to substantial change within the system might occur.

## 4. Reflections

Among co-producers from outside [ANONYMISED], the change of focus from child obesity to child health, and evidence that their input was recognized, was highly valued. Many reported feeling that they personally benefitted from participating in the project. Some indicated that they are very keen to take part in further research, and some that they had heard perspectives which may lead to changes in their professional practice. Generally, co-producers across all three workshops indicated that the model more accurately reflected the complexity of childhood health than simpler models that focus on obesity status (usually operationalised as BMI) as the outcome.

There was a strong feeling from people with lived experience that research and services repeat the same mistakes repeatedly, embedding stigma and focus on individuals rather than the wider context. One participant had come into the project expecting this to be the case, was delighted to find otherwise, and thrilled that their contributions were having a meaningful impact. However, they felt the final result did not go far enough in making progress.

There was also some feedback from stakeholders that the logic model was perhaps too complex to be usable by some of its intended audience. The participants also noted visual elements that could be refined to make the balance of the different factors clearer (for example, one participant noted that some of the levels had larger boxes than others, which could give a misleading impression about their relative importance).

## 4.1 Reflections from the [ANONYMISED]-based co-producers

Co-production inputs changed the focus and our collective understanding from a potentially stigmatising focus on obesity towards a more holistic understanding of childhood health. This focus also allowed for greater consideration of the social determinants of health and the broader macro-level factors that influence children’s health.

As many of us reflected, co-production gave us legitimacy in stepping away from academic conventions that could otherwise be harmful. We felt empowered by our co-producers to take on a perspective which was in alignment with our knowledge of evidence in the area and of our values and experiences as people.

Co-producing the model also underscored to us to the stigmatising capacity of labels that indicate health and ill-health. While we felt as a team that we were making a radical departure from conventional academic practice in our choice of outcome, we also acknowledge that this departure did not go far enough for some.

We reflected that while the model was co-produced, we also worked with the advisory group in a way that mirrored co-production. We also recognised that co-production did challenge our usual ways of working and this transition was not always comfortable or easy to implement. For most of the [ANONYMISED]-based co-producers, this was our first (or near-first) experience of co-production. Co-production is a transformational way of creating research and it is perhaps unsurprising that this first project represents a learning curve.

However, its transformational nature is indisputable and highly valued across all involved, and we have all continued to embed co-production within subsequent projects and to embrace the steep learning curve each time.

## 5. Discussion

This study reveals the capacity of lay and public contributors to develop complex theories that can be the basis of later research. Our model contains a number of features that can be associated with intervention complexity, including its focus on theorising contextual features that may interact with intervention features ^9,25^ and a high number of upstream and distal factors, with individual factors broadly treated as mediators ^26^. Other forms of complex causal relationships were also represented within the model. For example, an example of conjunctural causation was recognised with the need to “make healthy food more convenient AND more affordable” within the model. Within the confines of the workshops, there was not sufficient time for further theorising about other forms of complex relationships, although this would have been a natural extension of the work undertaken.

### 5.1 The strengths of co-producing a logic model

While public involvement is recommended in the development of logic models (see for example ^6,26^), there were few examples at the time in which this research took place where public involvement had been documented, and particularly a description of how public involvement influenced the contents of the model. Since then, a wider body of literature has emerged that documents that public involvement is possible and beneficial to the creation of theory. These studies emphasise the role of working with public contributors in strengthening understanding between researchers and stakeholders (for example ^27,28^), through developing a common language between contributors with different perspectives (for example ^27,29^) that can help to create research that represents a deeper understanding of a phenomenon that is perceived to be more relevant and applicable ^28-31^.

In previous exercises, academic teams have attempted to position themselves as a neutral party that plays a role in facilitating the smoothing of relations between other stakeholders (e.g., between marginalised communities and empowered others such as service providers and policy-makers) (see ^32^). In our case, [ANONYMISED]-based co-producers were the historically empowered stakeholder, representing the party that had been setting the discourse around research on childhood health. We adopted an enquiring and responsive approach, rather than a neutral approach.

### 5.2 The challenges of co-producing a logic model

Challenges and difficulties in public involvement in logic models are highlighted in the literature, including difficulties in reconciling different perspectives, uneven participation among group members, and a poor connection between the logic model and its intended use ^30^. This latter factor is a particular risk for our example, where there exists a risk that the richness of the model is not adequately reflected within published literature. However, we do not view this as a deficiency of the logic model or the theorising *per se*, but a limitation of published literature, a great deal of which has not have been conducted with meaningful public involvement.

Moreover, some professional participants at the third workshop noted the challenges of using or engaging with such a complex model. Further, when processes and services are in place, and expectations are entrenched, it may be hard to shift the emphasis on BMI to a broader set of outcome measures that do not link to a clearcut clinical definition of child health. Similarly, the model also presents a challenge around how and where to intervene. Organisations such as school typically have little leverage in changing higher-level factors in the system (e.g. at the sociopolitical level). Nevertheless, the influence of these system-wide factors on children’s health remains important to acknowledge.

Finally, some literature questions the benefits of deep co-production in research due to its time and resource consuming nature (for example ^33^). There is no doubt that setting out to co-produce a logic model was a complex undertaking. However, we also reflect that co-production is about conducting better research through working in an inclusive way, and that working inclusively takes time.

Where we do see limitations to co-production, these are more in the research systems – including funding and research ethics systems – that can hinder the enactment of co-production and serve to devalue the work of people with lived experience as equal or even relevant. The findings here clearly attest to the multifarious and transformational benefits of co-production.

### 5.3 The contribution of the new model

The strengths of our way of working and the model itself are in its transformational lens on how we should conceptualise childhood obesity. This is directly attributable to the influence of public involvement and stands apart from the earlier Foresight Obesity map ^16^.

Shortcomings in the development of the Foresight map have also been highlighted by others, who have also responded through developing their own obesity systems map. McGlashan and colleagues ^33^ developed a systems map on childhood obesity informed by community representatives from schools, health services, sporting clubs and local government. They found that variables specific to the school environment were not included in the Foresight map and the authors conclude that their community-driven map was more closely aligned with a more locally relevant and feasible intervention strategy ^33^.

Luna Pinzon and colleagues ^34^also differed in who informed their adolescent obesity systems map (a causal loop diagram). They took a multi-actor perspective including academic researchers, adolescents, and local stakeholders, although these groups worked in parallel and not directly together. Drawing causal loop diagrams based on these different perspectives, they found overall that the researchers’ and stakeholders’ perspectives contributed more to how the system works whereas integrating the lived experience of the adolescents enriched the insights on how they, the adolescent, interacted with that environment ^34^. Both this and the McGlashan et al. examples retained the focus on obesity, although in the case of Luna Pinzon et al ^34^, this was conceptualised as obesity-related behaviours (namely adolescents’ dietary behaviour, physical activity, sedentary behaviour, and sleep).

Where our own model contributes to the literature in this area is in representing a synthesis of the views of stakeholders who co-produced the logic model together. The product of this co-production sends clear signals that (i) the historical focus on childhood obesity – understood through a single clinical measure (BMI) - is stigmatising and can belie the complexity of factors that contribute to poor health in childhood (represented alternatively in our model through the focus on HEPAM); (ii) many of the factors that are theorised to contribute to poorer health, and that influence the capacity of schools to improve child health, occur at the policy level; and (iii) theories co-produced with a broad swathe of co-producers emphasise the role of the social determinants of health and serve as a useful challenge to those that focus heavily on narratives of personal responsibility and individual determinism in children’s health.

## 6. Conclusions

Our attempt at co-production was a transformational experience. Our work underscores that childhood obesity is a clinical diagnosis for a state of poor child health that has stigmatising consequences. Working as a large group with diverse experiences and perspectives challenged orthodox academic conventions that instinctively gravitate towards medicalised understandings of health, and which tend to look for individual-level, proximal, or downstream causes of poorer health. Among the [ANONYMISED]-based co-production team, despite our own values and research experiences, we have reflected that without co-production we may have repeated these same conventional explanations, thereby continuing to embed stigma and focus on factors reflecting personal responsibility rather than the wider context.

## Data Availability

The data that support the findings of this study are available in the main body or are freely available here: https://sites.google.com/view/cephi-project/logic-model. No other data were collected or are available due to privacy or ethical restrictions.

## Acknowledgments

Thank you to all those who contributed their time, expertise and experience to co-create the logic model with us, without whom this project would look very different. The research was funded by National Institute of Health Research under the Public Health Research stream (PHR Project: NIHR133736 - Handling complexity in evidence from systematic reviews and meta-analyses of public health intervention).

